# Characterising the persistence of RT-PCR positivity and incidence in a community survey of SARS-CoV-2

**DOI:** 10.1101/2021.08.12.21261987

**Authors:** Oliver Eales, Caroline E. Walters, Haowei Wang, David Haw, Kylie E. C. Ainslie, Christina Atchison, Andrew J. Page, Sophie J. Prosolek, Alexander J. Trotter, Thanh Le Viet, Nabil-Fareed Alikhan, Leigh M Jackson, Catherine Ludden, The COVID-19 Genomics UK (COG-UK) Consortium, Deborah Ashby, Christl A. Donnelly, Graham Cooke, Wendy Barclay, Helen Ward, Ara Darzi, Paul Elliott, Steven Riley

## Abstract

**Background:** Community surveys of SARS-CoV-2 RT-PCR swab-positivity provide prevalence estimates largely unaffected by biases from who presents for routine case testing. The REal-time Assessment of Community Transmission-1 (REACT-1) has estimated swab-positivity approximately monthly since May 2020 in England from RT-PCR testing of self-administered throat and nose swabs in random non-overlapping cross-sectional community samples. Estimating infection incidence from swab-positivity requires an understanding of the persistence of RT-PCR swab positivity in the community.

**Methods:** During round 8 of REACT-1 from 6 January to 22 January 2021, of the 2,282 participants who tested RT-PCR positive, we recruited 896 (39%) from whom we collected up to two additional swabs for RT-PCR approximately 6 and 9 days after the initial swab. We estimated sensitivity and duration of positivity using an exponential model of positivity decay, for all participants and for subsets by initial N-gene cycle threshold (Ct) value, symptom status, lineage and age. Estimates of infection incidence were obtained for the entire duration of the REACT-1 study using P-splines.

**Results:** We estimated the overall sensitivity of REACT-1 to detect virus on a single swab as 0.79 (0.77, 0.81) and median duration of positivity following a positive test as 9.7 (8.9, 10.6) days. We found greater median duration of positivity where there was a low N-gene Ct value, in those exhibiting symptoms, or for infection with the Alpha variant. The estimated proportion of positive individuals detected on first swab, *P*_0_, was found to be higher for those with an initially low N-gene Ct value and those who were pre-symptomatic. When compared to swab-positivity, estimates of infection incidence over the duration of REACT-1 included sharper features with evident transient increases around the time of key changes in social distancing measures.

**Discussion:** Home self-swabbing for RT-PCR based on a single swab, as implemented in REACT-1, has high overall sensitivity. However, participants’ time-since-infection, symptom status and viral lineage affect the probability of detection and the duration of positivity. These results validate previous efforts to estimate incidence of SARS-CoV-2 from swab-positivity data, and provide a reliable means to obtain community infection estimates to inform policy response.

## Introduction

Symptom-initiated community testing for SARS-CoV-2 is a vital public health intervention that enables the isolation of known positives and the quarantining of close contacts [1]. However, the trend of cases obtained from routine surveillance is often unreliable because of capacity issues and changing propensity to seek tests [2]. Therefore, data from representative community studies of swab positivity such as the REal-time Assessment of Community Transmission-1 (REACT-1) study [3] are used to infer prevalence of swab-positivity in order to maintain situational awareness during periods when routine testing is less reliable. In order to infer infection incidence from these data, estimates are required of test sensitivity and the duration that people continue to test positive, but these depend heavily on factors that will vary between settings and study designs. For example, estimates of sensitivity of RT-PCR to detect SARS-CoV-2 have been made using samples taken by health care professionals from hospitalized patients [4,5], and from small non-representative groups [6]. Also, the specific criteria used to declare results from RT-PCR assays as positive, such as use of multiple gene targets and limits of cycle threshold (Ct) values, vary from laboratory to laboratory.

REACT-1 is tracking the spread of SARS-CoV-2 in England over time at national and regional scales and in different socio-demographic groups [3], based on self-administered throat and nose swabs obtained from non-overlapping random samples of the population [7]. However, estimates of daily incidence during the pandemic have relied on unvalidated assumptions concerning RT-PCR sensitivity and the duration of RT-PCR positivity.

Here, in a substudy of REACT-1 round 8 (6 January 2021 - 22 January 2021) [8] we asked participants who tested positive to obtain two additional swabs approximately 5 days apart. In this way we aimed to estimate the study-specific sensitivity of RT-PCR based on a single swab, and the average duration for which individuals remained positive, allowing us to provide daily incidence estimates over time.

## Results

Of the 2,282 individuals testing positive in round 8, 896 (39%) agreed to take part in this sub-study, of whom 874 (98%) had more than one successful RT-PCR test with valid date information (662 with three tests, 212 with two tests). The median interval between the first positive test and second test was 6 days, and it was 9 days between the first and third test, with the largest delay between the first and last test being 17 days. Of the 874 participants with at least one valid additional test: 323 (37%) were positive on all additional tests (237 with two additional tests and 86 with one additional test); 412 (47%) were negative on all additional tests (286 with two additional tests and 126 with one additional test); and the remainder had a mix of positive and negative results on additional tests (Figure 1A).

**Figure 1.**
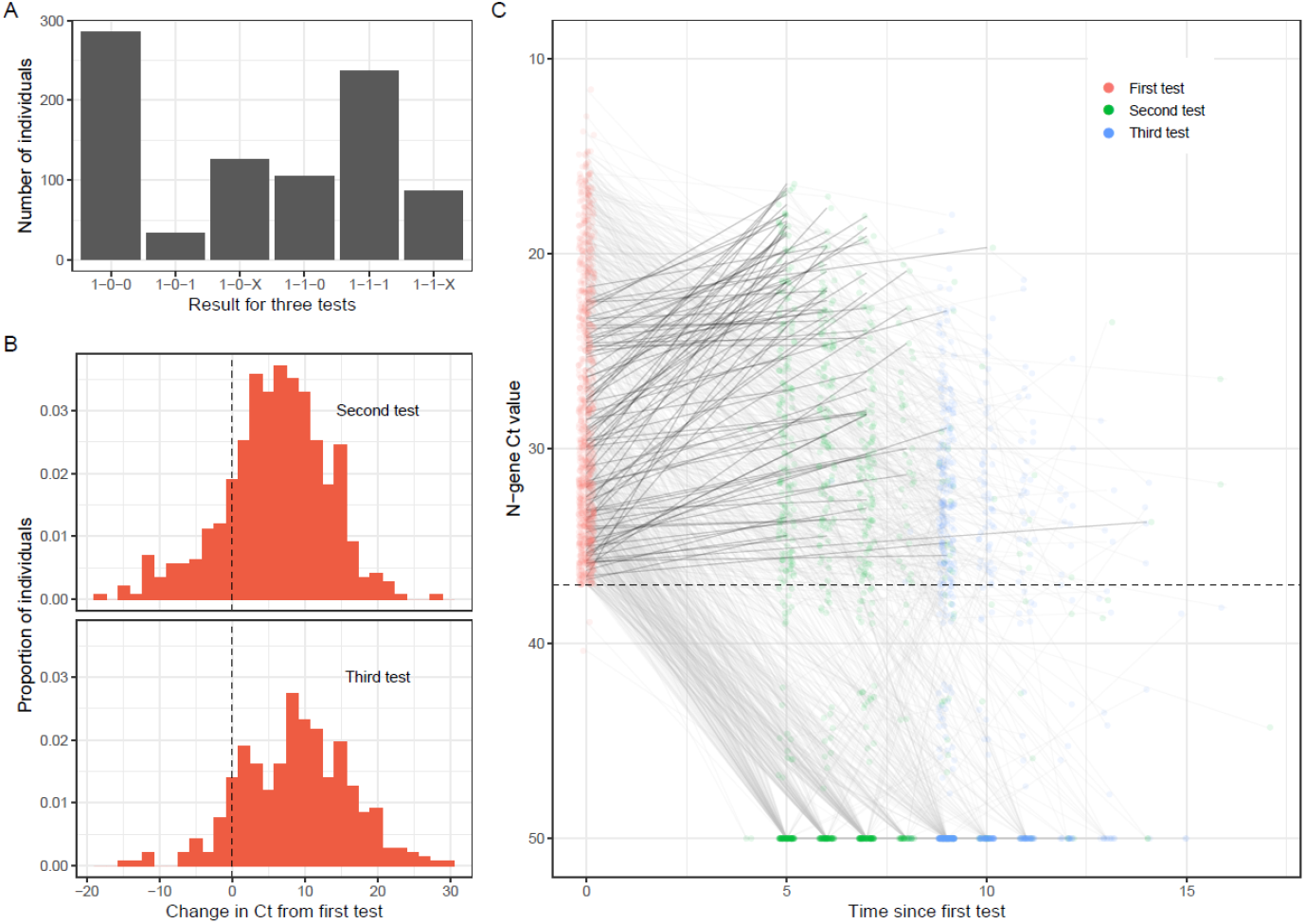
Patterns in positivity and N-gene Ct value over time. (A) The number of individuals with each combination of test results. “1” represents a positive test result, “0” represents a negative test result, “X” that one test was undetermined and so excluded from the analysis. The ordering represents the order in which test results were obtained. (B) Distribution of changes in Ct value between the first (positive) test and following tests. Only tests with an accurate estimate of N-gene Ct value are included; those tests in which the N-gene Ct was not measured successfully which were included with an N-gene Ct value of 50 (the limit of detection) in panel A have not been included. (C) N-gene Ct value measured in repeat tests over time. Points show the data for the first measurement (red), second measurement (green) and third measurement (blue) of N-gene Ct value against time (jittered). The lines connect each individual’s repeat measurements; decreases in Ct (dark grey) between first measurement and second measurement have been highlighted through a darker coloring. Points with a N-gene Ct value of 50 did not have any virus detected and so have been placed at the limiting value of the test.

We developed a statistical model of positivity to describe the probability of participants testing positive as a function of time since the first positive sample (see Methods). We fit an exponential decay in the probability of being positive, with a decay rate and initial proportion of positive individuals detected, *P*_0_, estimated from the data (Figure 2). For all participants, we estimated *P*_0_ as 0.79 (0.77, 0.81) and daily decay rate 0.071 (0.065, 0.078) corresponding to a median duration positive of 9.7 (8.9, 10.6) days, or a mean duration positive of 14.0 (12.9, 15.4) days (under the assumption of an exponential trend, Table 1).

**Table 1.** Parameter estimates for all subsets of the data.

| Data | Subset | N | Model | Parameter 1 (k) | Parameter 2 (P<sub>0</sub>) | Parameter 3 (plateau duration)* | Median duration positive | Mean duration positive |
| --- | --- | --- | --- | --- | --- | --- | --- | --- |
| All | All | 874 | Model 1 | 0.071 ( 0.065 , 0.078 ) | 0.79 ( 0.77 , 0.81 ) |  | 9.7 ( 8.9 , 10.6 ) | 14.0 ( 12.9 , 15.4 ) |
| Ct value | Ct ≤ 24.5 | 287 | Model 1 | 0.026 ( 0.017 , 0.037 ) | 0.91 ( 0.86 , 0.95 ) |  | 26.4 ( 18.9 , 40.8 ) | 38.1 ( 27.3 , 58.9 ) |
|  |  |  | Model 2** | 0.064 ( 0.045 , 0.084 ) | 0.95 ( 0.91 , 0.98 ) | 3.97 ( 3.00 , 4.52 ) | 14.8 ( 12.4 , 18.9 ) | 19.6 ( 16.1 , 25.8 ) |
|  | 24.5 < Ct ≤ 32.5 | 302 | Model 1 | 0.088 ( 0.071 , 0.105 ) | 0.82 ( 0.74 , 0.89 ) |  | 7.9 ( 6.6 , 9.8 ) | 11.4 ( 9.5 , 14.1 ) |
|  | Ct > 32.5 | 285 | Model 1 | 0.143 ( 0.105 , 0.171 ) | 0.52 ( 0.39 , 0.61 ) |  | 4.9 ( 4.0 , 6.6 ) | 7.0 ( 5.8 , 9.5 ) |
| Symptoms<br>(in month<br>before first<br>test) | Any symptoms | 549 | Model 1 | 0.057 ( 0.048 , 0.065 ) | 0.81 ( 0.77 , 0.83 ) |  | 12.2 ( 10.7 , 14.4 ) | 17.7 ( 15.5 , 20.7 ) |
|  | Classic<br>COVID symptoms** | 391 | Model 1 | 0.053 ( 0.042 , 0.063 ) | 0.79 ( 0.73 , 0.83 ) |  | 13.1 ( 11.0 , 16.6 ) | 18.9 ( 15.9 , 23.9 ) |
|  | No symptoms*** | 325 | Model 1 | 0.132 ( 0.114 , 0.151 ) | 0.94 ( 0.88 , 0.98 ) |  | 5.3 ( 4.6 , 6.1 ) | 7.6 ( 6.6 , 8.8 ) |
|  | Pre-symptomatic | 107 | Model 1 | 0.061 ( 0.045 , 0.081 ) | 0.99 ( 0.95 , 1.00 ) |  | 11.3 ( 8.5 , 15.5 ) | 16.3 ( 12.3 , 22.3 ) |
|  | Asymptomatic | 218 | Model 1 | 0.182 ( 0.145 , 0.219 ) | 0.81 ( 0.62 , 0.93 ) |  | 3.8 ( 3.2 , 4.8 ) | 5.5 ( 4.6 , 6.9 ) |
| Lineage | Alpha variant | 368 | Model 1 | 0.034 ( 0.025 , 0.043 ) | 0.91 ( 0.86 , 0.95 ) |  | 20.4 ( 16.1 , 27.4 ) | 29.5 ( 23.2 , 39.5 ) |
|  |  |  | Model 2** | 0.065 ( 0.047 , 0.085 ) | 0.94 ( 0.90 , 0.97 ) | 3.43 ( 2.16 , 4.16 ) | 14.0 ( 12.0 , 17.5 ) | 18.7 ( 15.6 , 24.1 ) |
|  | Wildtype | 75 | Model 1 | 0.085 ( 0.056 , 0.119 ) | 0.87 ( 0.72 , 0.96 ) |  | 8.1 ( 5.8 , 12.4 ) | 11.7 ( 8.4 , 17.9 ) |
| Age | 18-40 | 256 | Model 1 | 0.076 ( 0.059 , 0.093 ) | 0.85 ( 0.77 , 0.91 ) |  | 9.2 ( 7.5 , 11.7 ) | 13.2 ( 10.8 , 16.9 ) |
|  | 41-59 | 359 | Model 1 | 0.066 ( 0.056 , 0.076 ) | 0.79 ( 0.74 , 0.82 ) |  | 10.6 ( 9.2 , 12.5 ) | 15.2 ( 13.2 , 18.0 ) |
|  | 60+ | 259 | Model 1 | 0.103 ( 0.085 , 0.122 ) | 0.90 ( 0.83 , 0.95 ) |  | 6.8 ( 5.7 , 8.1 ) | 9.7 ( 8.2 , 11.7 ) |

Parameters shown are the exponential decay rate (k), initial proportion of positive individuals detected (Sensitivity proxy) and for Model 2 the additional time delay parameter.
Values estimated from parameters are median duration positive, calculated as $\log(2)/k$ with the time delay added on for Model 2, and mean duration positive, calculated as $1/k$ with the time delay added on for Model 2
\* Model 2 is only presented for the subsets of data where it was found to be a better fit (see Methods)
\*\* Loss or change of sense of smell, loss or change of sense of taste, new persistent cough, fever
\*\*\* Those reporting no symptoms have been subset into those that report symptoms in the past month during their subsequent tests (Pre-symptomatic) and those that do not (Asymptomatic)

**Figure 2:**
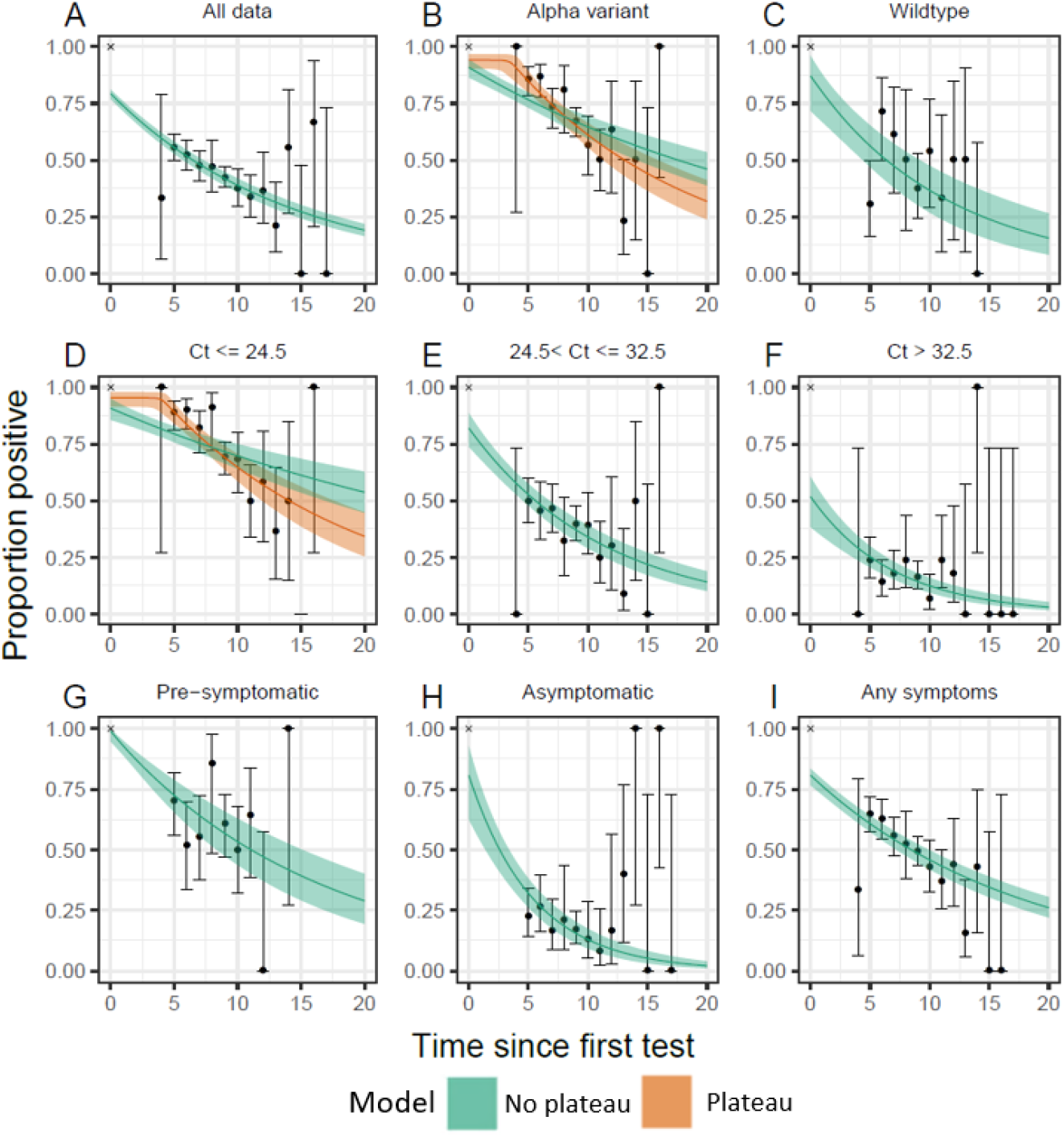
Best-fitting exponential decay model with no plateau (dark green line) and 95% confidence interval (green shaded region). Best-fitting exponential decay model with a plateau (dark orange) and 95% confidence interval (orange shaded region) are included for the subsets of data where the model was preferred. Data (black points) and 95% Binomial confidence intervals (black error bars) are included for the proportion of tests that were positive. (A) Model fit to all available data. (B,C) Model fits to subsets of the data based on the determined lineage (Alpha, wildtype). (D,E,F) Model fits to subsets of the data based on N-gene Ct value of the first test (Ct<=24.5, 24.5<Ct<=32.5, Ct>32.5). (G,H,I) Model fits to subsets of the data based on symptom status: those reporting any symptoms in the month prior to the first test (Any symptoms), those reporting no symptoms prior to all their tests (Asymptomatic) and those reporting no symptoms prior to their first test but symptoms prior to subsequent tests (Pre-symptomatic). Note that the models are not fit to the data as presented, but to the exact ordering of test outcomes for each individual.

### Translating swab-positivity to incidence

With the estimates for *P*_0_ (equivalent to the sensitivity of the study) and mean duration positive (Table 1) we converted previous estimates of weighted swab-positivity for each round of REACT-1 into estimates of average daily incidence (Table 2). For round 8 (6 January 2021 - 22 January 2021), in which swab-positivity was highest of all rounds of REACT-1 to date, the swab positivity of 1.57% (1.49%, 1.66%) corresponded to an average daily incidence of 0.141% (0.126%, 0.158%), which translates to an average of 79,736 (71083, 89519) daily infections given the population of England. For round 13 (24 June 2021 - 12 July 2021), the most recent round, the swab positivity of 0.63% (0.57%, 0.69%) corresponded to an average daily incidence of 0.057% (0.049%, 0.065%), or 32,007 (27823, 36071) daily infections.

**Table 2.** Weighted swab-positivity, daily incidence rate and incidence for each round of REACT-1 with 95% confidence intervals.

| Round | First Sample | Last Sample | Tested swabs | Positive swabs | Weighted swab positivity | Daily incidence rate | Daily incidence (N*) |
| --- | --- | --- | --- | --- | --- | --- | --- |
| 1 | 2020-05-01 | 2020-06-01 | 120,620 | 159 | 0.16% ( 0.13% , 0.19% ) | 0.014% ( 0.012% , 0.018% ) | 8143 ( 6503 , 9953 ) |
| 2 | 2020-06-19 | 2020-07-07 | 159,199 | 123 | 0.09% ( 0.07% , 0.11% ) | 0.008% ( 0.006% , 0.010% ) | 4575 ( 3506 , 5712 ) |
| 3 | 2020-07-24 | 2020-08-11 | 162,821 | 54 | 0.04% ( 0.03% , 0.05% ) | 0.004% ( 0.003% , 0.005% ) | 2036 ( 1504 , 2601 ) |
| 4 | 2020-08-20 | 2020-09-08 | 154,325 | 137 | 0.13% ( 0.10% , 0.15% ) | 0.012% ( 0.009% , 0.014% ) | 6616 ( 5237 , 8087 ) |
| 5 | 2020-09-18 | 2020-10-05 | 174,949 | 824 | 0.60% ( 0.55% , 0.71% ) | 0.054% ( 0.045% , 0.063% ) | 30480 ( 25674 , 35853 ) |
| 6 | 2020-10-16 | 2020-11-02 | 160,175 | 1,732 | 1.30% ( 1.21% , 1.39% ) | 0.117% ( 0.103% , 0.132% ) | 65994 ( 58360 , 74646 ) |
| 7 | 2020-11-13 | 2020-12-03 | 168,181 | 1,299 | 0.94% ( 0.87% , 1.01% ) | 0.084% ( 0.074% , 0.096% ) | 47728 ( 42073 , 54118 ) |
| 8 | 2021-01-06 | 2021-01-22 | 167,642 | 2,282 | 1.57% ( 1.49% , 1.66% ) | 0.141% ( 0.126% , 0.158% ) | 79736 ( 71083 , 89519 ) |
| 9 | 2021-02-04 | 2021-02-23 | 165,456 | 689 | 0.49% ( 0.44% , 0.55% ) | 0.044% ( 0.038% , 0.051% ) | 24882 ( 21319 , 28841 ) |
| 10 | 2021-03-11 | 2021-03-30 | 140,844 | 227 | 0.20% ( 0.17% , 0.23% ) | 0.018% ( 0.015% , 0.021% ) | 10179 ( 8426 , 12102 ) |
| 11 | 2021-04-15 | 2021-05-03 | 127,408 | 115 | 0.10% ( 0.08% , 0.13% ) | 0.009% ( 0.007% , 0.012% ) | 5084 ( 3766 , 6503 ) |
| 12 | 2021-05-20 | 2021-06-07 | 108,911 | 135 | 0.15% ( 0.12% , 0.18% ) | 0.014% ( 0.011% , 0.017% ) | 7634 ( 5994 , 9387 ) |
| 13 | 2021-06-24 | 2021-07-12 | 98,233 | 527 | 0.63% ( 0.57% , 0.69% ) | 0.057% ( 0.049% , 0.065% ) | 32007 ( 27823 , 36701 ) |

\*Number of people infected each day calculated using mid-2020 population estimates for England

Fitting a P-spline model to incidence and swab-positivity separately (see Methods) we find that patterns in incidence preceded changes in swab-positivity (Figure 3) and that features on the incidence time series were sharper than the corresponding features of the swab-positivity time series. For example, there was a more pronounced peak in mid-October in incidence compared with swab-positivity, which fell sharply until mid-November. From mid-November onwards incidence increased and appeared to begin decreasing in early January, though with no data from REACT-1 for December there was considerable uncertainty in the estimates for this period. From early May to beginning of July 2021, both incidence and swab-positivity increased exponentially. However, by the end of round 13 (12 July 2021) it appeared that incidence was no longer increasing, though with wide credible intervals on the P-spline.

**Figure 3.**
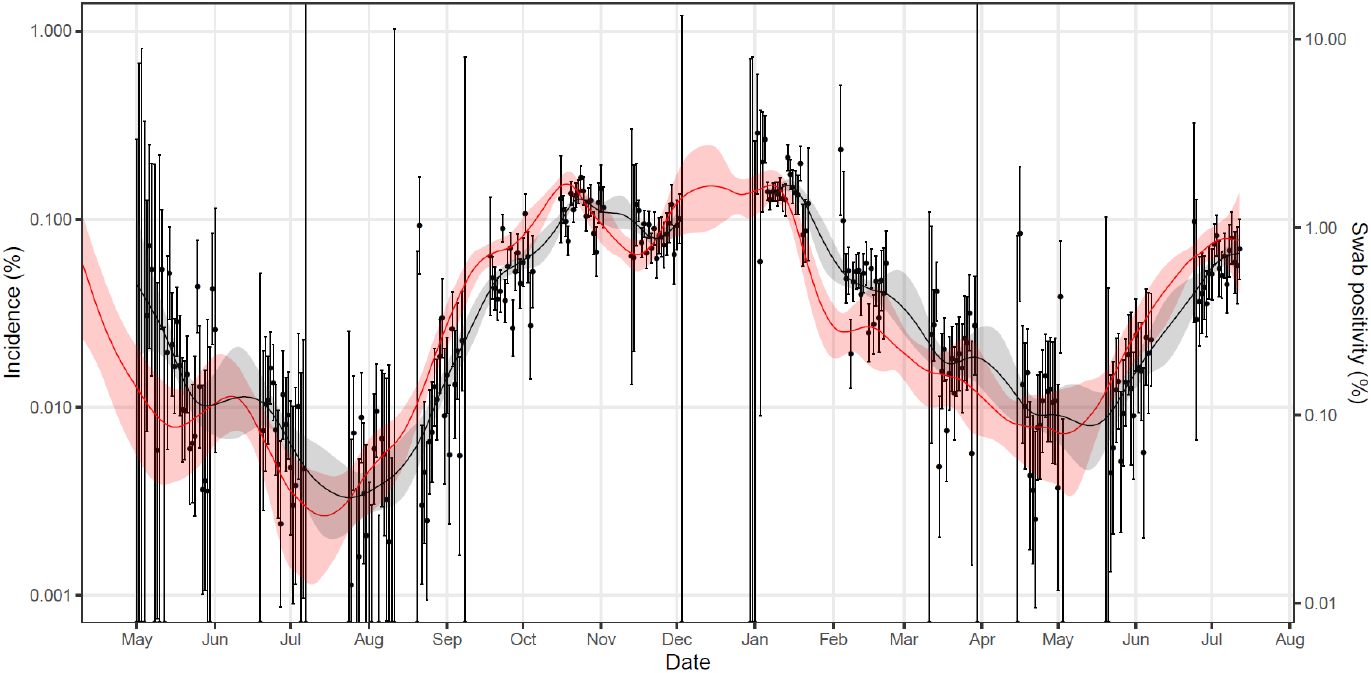
Comparison of inferred daily incidence and swab-positivity over 13 rounds of REACT-1 with a *log*_10_ y-axis. Modelled estimate of daily incidence (red line, left hand y-axis) with 95% credible interval (red shaded region) across all 13 rounds of REACT-1. Modelled estimate of daily swab positivity (black, right-hand y-axis) with 95% credible interval (grey shaded region) for all 13 rounds of REACT-1. Note that the model estimates are not shown for the period between rounds 7 and 8 (December) as there were no data to accurately capture the December peak in swab-positivity. Daily weighted observations (points) and 95% Binomial confidence intervals (vertical lines) are also shown for swab positivity (right hand axis).

### Subgroup analyses

We found evidence of differences in the time course of positivity driven by a number of factors: Ct value, symptom status, lineage, and age (Figure 2, Table 1, Supplementary Figure 1). Also, we extended the statistical model of positivity to include a plateau before the start of the exponential decay and fit models with and without the plateau to subsets of the data selecting the best-fitting version (see Methods).

### Ct value

We found that the estimates of median duration of positivity and *P*_0_ were dependent on N-gene Ct value (Figure 4, Table 1). The estimate of *P*_0_ was highest at 0.95 (0.91, 0.98) for participants whose initial Ct value was in the lowest ∼1/3 (less than or equal to 24.5), indicating high sensitivity to detect strong positives, decreasing to 0.52 (0.39, 0.61) in those participants with a Ct value in the highest ∼1/3 (greater than 32.5). The estimated median duration of positivity was highest in those with a Ct <= 24.5 at 14.8 (12.4, 18.9) days decreasing to 4.9 (4.0, 6.6) days for those with a Ct greater than 32.5. The best-fitting model for those with a Ct value less than 24.5, included a 4.0 (3.0, 4.5) day plateau before the exponential decay (difference in expected log predictive density [ELPD] = 9.5).

**Figure 4:**
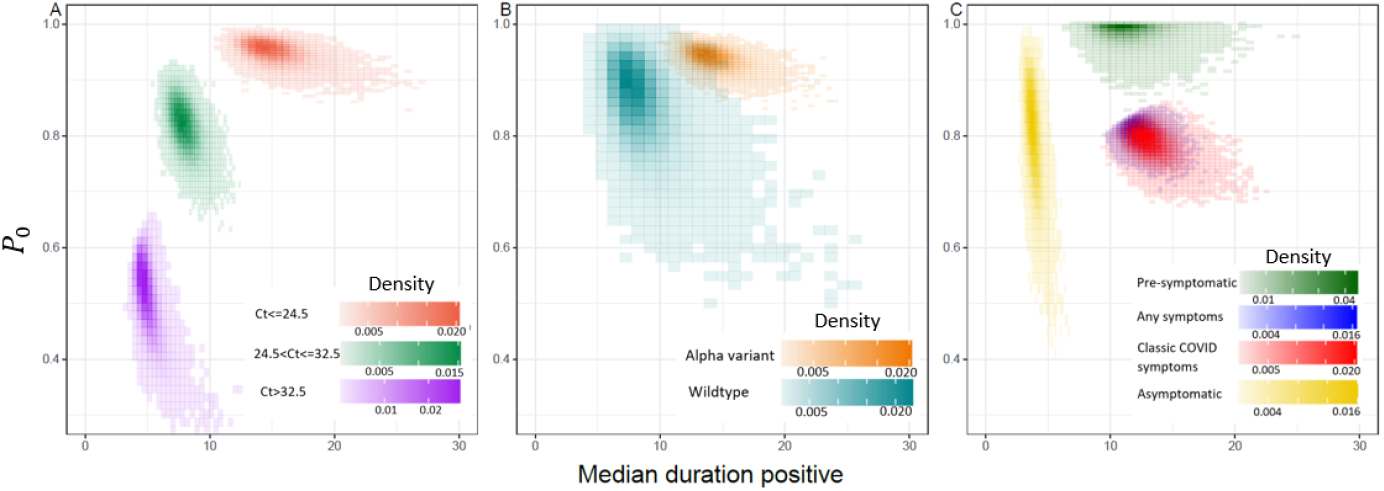
Posterior probability density for the initial proportion testing positive, *P*_0_, and the median duration of positivity. (A) Comparison of the posterior probability density for models fit to subsets of the data based on the N-gene Ct value of the first test (Ct<=24.5, 24.5<Ct<=32.5, Ct>32.5). (B) Comparison of the posterior probability density for models fit to subsets of the data based on determined lineage (Alpha, wildtype). (C) Comparison of the posterior probability density for models fit to subsets of the data based on symptoms status in the month prior to their first test (Pre-symptomatic, any symptoms, most predictive COVID-19 symptoms, asymptomatic). Note that for the subset of data in which the lineage was determined to be the Alpha variant, and for the subset of data in which N-gene Ct value was less than 24.5 this is the density from the exponential decay model with an initial plateau (Model 2).

In addition to positivity, we analysed Ct values across repeated tests for the same individuals (Figure 1). N-gene Ct values in the first (positive) test ranged from 11.6 to 40.4 with a mean of 28.0. In subsequent tests, overall, N-gene Ct values increased, though for a small proportion of individuals they decreased from the first test to subsequent tests. However, we were unable to estimate N-gene Ct values for 41% of second tests and 47% of third tests, either due to a negative test or because the Ct value was higher than the limit of reporting (N-gene Ct = 50).

### Symptom status

We found differences in the estimates of *P*_0_ and the median duration of positivity according to symptom status. Participants who reported any symptoms and those with the most predictive COVID-19 symptoms (loss or change of sense of smell, loss or change of sense of taste, new persistent cough, fever [9]) in the month prior to their first test had median durations of positivity of 12.2 (10.7, 14.4) days and 13.1 (11.0, 16.6) days respectively. In contrast, those with no reported symptoms in the month prior to their first test had a shorter median duration of positivity at 5.3 (4.6, 6.1) days (Table 1). Subsetting these individuals into those who reported any symptoms in subsequent tests (pre-symptomatics) and those who reported no symptoms (asymptomatics) also identified clear differences (Figure 4, Table 1). Pre-symptomatic individuals had an estimated median duration of positivity of 11.3 (8.5, 15.5) days, and for those initially reporting symptoms it was 12.2 (10.7, 14.4) days. Asymptomatic individuals had a shorter estimated median duration of positivity at 3.8 (3.2, 4.8) days. The estimated initial proportion of positives detected, *P*_0_, was highest in pre-symptomatic individuals at 0.99 (0.95, 1.00). In comparison with asymptomatic individuals, for those reporting any symptoms and those reporting the most predictive COVID-19 symptoms, estimates of *P*_0_ were similar at 0.81 (0.62, 0.93), 0.81 (0.77, 0.83) and 0.79 (0.73, 0.83) respectively.

### Lineage

We found greater estimated median duration of positivity in those infected with the Alpha variant (previously called B.1.1.7) at 14.0 (12.0, 17.5) days versus 8.1 (5.8, 12.4) days in those infected with wildtype, although with overlapping confidence intervals (Figure 4, Table 1). We also fit models to random samples of individuals infected with Alpha variant, selected so that the proportion of individuals with initial N-gene Ct value <=24.5, 24.5<Ct<=32.5 and >32.5 was the same as for those infected with wildtype; this led to a slight reduction in the estimated median duration of positivity for Alpha variant, but it was still greater than the corresponding value for those infected with wildtype (Supplementary Table 1). The best-fitting model for those infected with the Alpha variant, included a 3.4 (2.2, 4.2) day plateau before the exponential decay (difference in ELPD = 5.4).

### Age

There were small differences in the estimates of *P*_0_ and the median duration positive between those aged 41-59 years and those aged 60 years and above, but no significant pairwise differences between those aged 18-40 years and these groups (Table 1).

## Discussion

We estimate the overall sensitivity of the study based on a single swab to be ∼79%, rising to 95% for strong positives, demonstrating why the RT-PCR remains the gold standard in testing for the presence of SARS-CoV-2. Additionally we characterised the median and mean duration that an individual remains positive after an initial positive test in REACT-1 at ∼10 and ∼14 days respectively (assuming a continued exponential decay beyond the follow-up period of our study), comparable to and validating previous estimates [10].

With both sensitivity and duration of positivity well-characterised we were able to convert our previous estimates of swab-positivity into incidence allowing estimates of new daily infections across the whole study period of REACT-1. This allowed us to assess the effects of various non-pharmaceutical interventions by comparing the patterns in incidence with the timing of changes in COVID-19 restrictions. For example, a fall in incidence was seen from mid-October 2020 suggesting that behaviour may have changed before the formal start of the lockdown introduced on 3 November 2020, and sooner than would be implied from the swab-positivity data. This is consistent with findings from a previous study that reported a decrease in mobility in the last two weeks of October 2020 [11].

The model used in calculating daily incidence has a number of limitations. Firstly we assumed an exponential decay to describe the probability of a participant testing positive in our study, which would not capture non-exponential longer-term trends in the waning of positivity. This limitation is unavoidable because our maximum follow-up was17 days after the initial positive infection, while it is known that people may remain positive for much longer periods [12]. Secondly, in estimating incidence we have assumed that the parameter estimates obtained for the decay of positivity from our round 8 substudy are representative of the entire study duration. Subgroup analysis showed there were differences in sensitivity and duration of positivity in individuals based on initial N-gene Ct value, symptomatology and viral lineage. With N-gene Ct value highly dependent on the contemporaneous growth rate of the pandemic [13], the distribution of Ct values may vary between rounds. Furthermore, lineages responsible for infections changed over the study period with the emergence of the Alpha variant in late 2020 [14], and the Delta variant in April 2021 [15]. We were unable to use lineage specific parameter estimates in estimates of infection incidence as lineage was only determined (via viral sequencing) in samples with lower Ct values (<34).

The median and mean durations of positivity we report do not directly inform isolation and quarantine policy. For example, the shorter duration of positivity we estimate for asymptomatic positives does not immediately suggest a shorter duration of isolation for asymptomatic contacts, despite the persistence of RT-PCR-positivity likely indicating some level of continued infectiousness: from our results, we cannot estimate the time at which symptoms occur in the 1/3 of those who initially did not report symptoms. Also, isolation and quarantine policies often reflect upper limits for durations of positivity, such as the 90th, 95th or even 97.5th percentiles [16], and must also take into account practical and logistical constraints on their implementation.

In subgroup analyses, as noted, we obtained higher sensitivity estimates for those infections with a lower Ct value suggesting that the test is more sensitive for stronger positives predominantly indicating recent infections (Ct is lowest at around ∼3 days [10]). Furthermore, estimated sensitivity was highest in presymptomatic individuals, whereas it was comparable between asymptomatic and symptomatic individuals, again suggesting that RT-PCR testing is highly effective at detecting early stage infections. Similar to a previous analysis [10] we found that median duration of positivity was lower in asymptomatic individuals, compared to those exhibiting any symptoms or one or more of the most predictive COVID-19 symptoms. Previous estimates of the proportion of infections that are asymptomatic, based on RT-PCR testing, may therefore have been underestimated due to their shorter duration. Finally we indicate that Alpha variant infections may have a greater median duration of positivity than wildtype, similar to other work [6], though with wide credible intervals. With the recent emergence of the Delta variant [15] in England, characterising any possible changes in RT-PCR test sensitivity and duration of positivity will be necessary to estimate infection incidence of Delta from swab-positivity data.

Our study provides similar data on Ct values to previous studies [6,10] that have estimated the duration of the proliferation and clearance stage of the virus. However, due to a lower rate of sampling in our study we are unable to estimate these parameters. The median duration of 6 days between the first and second test is greater than the estimated proliferation stage of ∼3 days. Additionally as recruitment to our study was based upon an initial positive result, and we only had swab results up to a maximum of 17 days post-first swab, we were particularly misplaced in estimating the course of the virus, as we will not have captured individuals at both early and late stages of infection.

As well as estimating test sensitivity and duration of positivity for the REACT-1 study, our results identify factors that will drive sensitivity and duration of positivity for community-based sampling more generally. Given likely reductions in many populations in the provision of routine community testing and in people’s propensity to seek tests, representative community PCR-based surveys similar to REACT-1 can continue to provide valuable situational awareness during periods of rapid changes in infection incidence.

## Methods

### Data

The methods for REACT-1 have been described before [7]. Since May 2020 there have been 13 rounds of data collected, approximately every month (except December 2020). For each round of the study a random cross-section of the population of England ages 5 years and over is sent a letter inviting them to take part. The invitees are randomly selected at the lower tier local authority level (n=315) through the list of general practitioners patients held by the National Health Service (NHS) in England. Those participants that agree to take part are sent a self-administered swab test (parent/guardian administered for those aged 5 to 12 years old). They also answer a short questionnaire providing information on age and other demographics as well as details on any symptoms that they exhibited in the month prior to their test. Swabs are collected in dry sample tubes and transported via cold chain to the laboratory for RT-PCR testing. In REACT-1, we obtain a single swab from each participant and test for N-gene and E-gene targets. We define a sample as positive if it has a valid positive signal for both gene targets (regardless of Ct value) or if it is positive only for N-gene with a Ct value equal to or lower than 37 [7].

During round 8 (6 January 2021 - 22 January 2021) the study participants who tested positive were invited to take an additional two swab tests. For those who agreed, swabs were administered and collected as before.

### Lineage designation

During round 8 of the study RT-PCR positive samples with a low enough N-gene Ct value (N-gene Ct value <34) and a high enough volume underwent genomic sequencing (Public Health England Research Ethics Governance Group (reference: R&D NR0195)). The methods have been described before [17]. In short, viral RNA was amplified using the ARTIC protocol [18] and sequence libraries were then prepared using CoronaHIT [19]. Raw sequence data were then analysed using the ARTIC bioinformatic pipeline [20] and lineage designation was performed using the machine learning-based assignment algorithm PangoLEARN [21]. Not all sequences obtained were of a sufficient quality for a lineage to be designated, and samples in which less than 50% of bases were covered were also deemed to have no lineage designation. Due to the repeat measurements in round 8 we were able to determine the lineage of infection as long as one swab test returned a definitive result. When discordant lineages were designated for the same individual the most advanced lineage was selected (e.g. B.1.1.7 over B.1) where one measurement just reflected a lower quality call, or if they were truly discordant (B.1.1.7 and B.1.117 for example) no lineage was designated. For the purposes of this paper, lineage segregated analysis only included individuals infected with the B.1.1.7 lineage (Alpha variant) or the wildtype (any lineage not a variant of concern or variant under investigation [22])

### Decaying positivity model

We model the probability of a positive test t days after an initial positive test. The models contain two components. The first is the initial proportion of positive individuals detected, *P*_0_, equivalent to the sensitivity of RT-PCR obtained from a single swab. The second component is the probability of an individual being positive after t days given they were positive at time t=0. We allow it to take two forms. The first (Model 1) is a simple exponential:

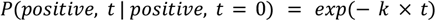

where k is the exponential decay rate. The second (Model 2) includes an extra parameter, τ, that introduces a plateau of duration τ before the exponential decay occurs:

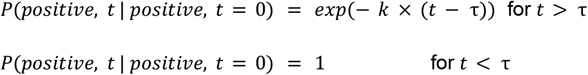

The Bayesian models are fit using a No-U-Turns Sampler [23] in Stan [24] with 10000 iterations and a burn-in of 500. Four chains are run for each model to assess whether there has been successful convergence. Writing *P*(*positive, t* | *positive, t* = 0) as *P*(*p, t*) for ease of notation the likelihood of the model for each possible individual outcome is:

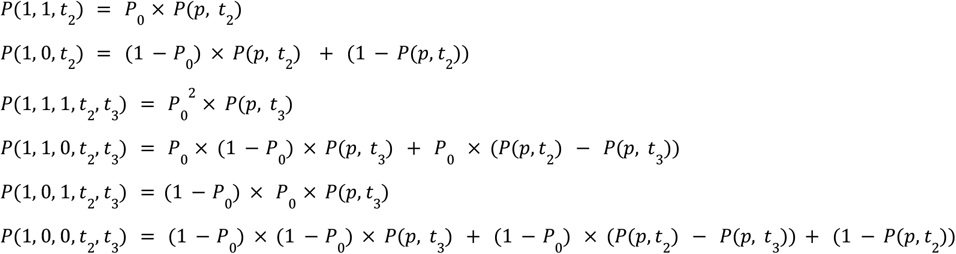

Where the probability on the left hand side denotes the outcome of all tests (two or three) with 1 representing a positive test and 0 representing a negative test. The times *t*_2_ and *t*_3_ are the number of days from the first test to the second test and third test respectively.

Model comparison between Model 1 and Model 2 was done by performing Pareto smoothed importance-sampling leave-one-out cross-validation (PSIS-LOO) [25]. Estimates of the ELPD and its standard error were calculated for both models and the more complex model (Model 2) was determined to be preferred if the value of the ELPD was greater than the ELPD of the simple model (Model 1).

Posterior samples of model fits were used to estimate the median and mean duration of positivity. Model 1 is simply an exponential distribution, for which the median value is given by *log*_*e*_ (2)/*k*, and the mean value is given by 1/*k*. For Model 2 the median and mean values are the same as for Model 1 but with the addition of the duration of the plateau.

### Converting swab-positivity to incidence

Estimates of the weighted swab-positivity for all 13 rounds of REACT-1 have previously been calculated [26]. Weighted swab-positivity is converted to average daily incidence by dividing by the sensitivity of the study (*P*_0_ from the decaying positivity model fit to all data), and by the mean duration of positivity (estimated from the decaying positivity model). The daily infection incidence is converted to an estimate for the number of daily infections in England using mid-2020 population estimates [27]. Estimates of average daily infection incidence are calculated for the entire posterior distribution of *P*_0_ and the mean duration of positivity with weighted swab-positivity randomly sampled from a normal distribution, with mean value the central estimate and standard deviation the width of the 95% CI divided by 3.92. From this posterior the median and 95% confidence interval are estimated for average daily incidence.

### P-spline incidence model

We fit a Bayesian P-spline model of incidence to the daily swab-positivity data for all 13 rounds of REACT-1. The period of time spanning from 50 days prior to the first day of the study to the last day of the study is segmented into regularly sized knots of approximately 5 days, with a further three knots defined before and after this period (to remove edge effects). A system of 4th order basis-splines (B-splines) is then defined over these knots, and the model of infection incidence is the linear combination of these B-splines. Overfitting of the model is minimised through the inclusion of a second-order random-walk prior distribution on the coefficients of the B-splines, *b*_*i*_ = 2*b*_*i* − 1_ − *b*_*i* − 2_ + *u*_*i*_, where *b*_*i*_ is the coefficient for the *i*^*th*^ B-spline, and *u*_*i*_ is normally distributed with *u*_*i*_ ∼ *N*(0, ρ^2^). The first two B-spline coefficients, *b*_1_ and *b*_2_, are assumed to have a uniform prior distribution. This prior distribution penalizes changes in the first derivative of the P-spline. The parameter ρ controls the level of penalisations and is assumed to have an uninformative inverse gamma prior distribution, ρ ∼ *IG*(0. 001, 0. 001). The P-spline of daily incidence is then converted into an estimate for the daily swab-positivity through the equation

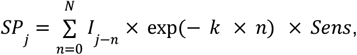

where *SP*_*j*_ is the swab-positivity on the *j*^*th*^ day, *I*_*j*−*n*_ is the incidence (from the P-spline) on the (*j* − *n*)^*th*^ day, *k* is the exponential decay rate and *Sens* is the sensitivity of the test. The exponential decay rate and sensitivity are taken to be the best estimates from the decaying positivity model. The summation should technically run from 0 to infinity but due to the need to compute it we take *N* to be 50 which was selected to be large enough that the exponential term in the model becomes negligible. By converting the daily incidence into a daily swab-positivity we are then able to fit our model to the daily data for swab-positivity that the study collects. The model is fit using a No-U-Turns Sampler (NUTS) [23] implemented in STAN [24].

### P-spline swab-positivity model

A Bayesian P-spline model of swab positivity was similarly fit. The entire study duration was segmented into regularly spaced knots of approximately 5 days, with extra knots extending beyond the period of the study to remove edge effects. Again a system of fourth order B-splines is defined over these knots and the P-spline model is a linear combination of these B-splines. Overfitting is as before minimised through the inclusion of a second-order random-walk prior distribution on the B-spline coefficients. As the P-spline is now modelling the swab-positivity there is no need for a transformation and the model can be fit directly to the daily swab-positivity data using a NUTS [23].

## Data Availability

These individual-level data are not yet available.

## Data availability

These individual-level data are not yet available.

## Acknowledgements

SR, CAD acknowledge support: MRC Centre for Global Infectious Disease Analysis, National Institute for Health Research (NIHR) Health Protection Research Unit (HPRU), Wellcome Trust (200861/Z/16/Z, 200187/Z/15/Z), and Centres for Disease Control and Prevention (US, U01CK0005-01-02). NFA was supported by the Quadram Institute Bioscience BBSRC funded Core Capability Grant (project number BB/CCG1860/1). GC is supported by an NIHR Professorship. HW acknowledges support from an NIHR Senior Investigator Award and the Wellcome Trust (205456/Z/16/Z). PE is Director of the MRC Centre for Environment and Health (MR/L01341X/1, MR/S019669/1). PE acknowledges support from Health Data Research UK (HDR UK); the NIHR Imperial Biomedical Research Centre; NIHR HPRUs in Chemical and Radiation Threats and Hazards, and Environmental Exposures and Health; the British Heart Foundation Centre for Research Excellence at Imperial College London (RE/18/4/34215); and the UK Dementia Research Institute at Imperial (MC_PC_17114). We thank The Huo Family Foundation for their support of our work on COVID-19. Quadram authors gratefully acknowledge the support of the Biotechnology and Biological Sciences Research Council (BBSRC); their research was funded by the BBSRC Institute Strategic Programme Microbes in the Food Chain BB/R012504/1 and its constituent project BBS/E/F/000PR10352. We thank members of the COVID-19 Genomics Consortium UK for their contributions to generating the genomic data used in this study. The COVID-19 Genomics UK (COG-UK) Consortium is supported by funding from the Medical Research Council (MRC) part of UK Research & Innovation (UKRI), the National Institute of Health Research (NIHR) and Genome Research Limited, operating as the Wellcome Sanger Institute.

We thank key collaborators on this work – Ipsos MORI: Kelly Beaver, Sam Clemens, Gary Welch, Nicholas Gilby, Kelly Ward and Kevin Pickering; Institute of Global Health Innovation at Imperial College: Gianluca Fontana, Sutha Satkunarajah, Didi Thompson and Lenny Naar; Molecular Diagnostic Unit, Imperial College London: Prof. Graham Taylor; North West London Pathology and Public Health England for help in calibration of the laboratory analyses; Patient Experience Research Centre at Imperial College and the REACT Public Advisory Panel; NHS Digital for access to the NHS register; and the Department of Health and Social Care for logistic support. SR acknowledges helpful discussion with attendees of meetings of the UK Government Office for Science (GO-Science) Scientific Pandemic Influenza – Modelling (SPI-M) committee.

## Additional information

Full list of COG-UK author’s names and affiliations are available in this [spreadsheet](https://drive.google.com/file/d/11Q3cnp2qGKMqcfrTKpp53jZFc7OdF0f7/view?usp=sharing).

**Supplementary Table 1:** Parameter estimates for the exponential decay model with no plateau (Model 1) and including a plateau (Model 2) for random subsets of individuals infected with the Alpha variant chosen to match the distribution of Ct values seen in individuals infected with wildtype.

| Lineage | Subset | N | Model | Parameter 1 (k) | Parameter 2 (P<sub>0</sub>) | Parameter 3 (plateau duration) | Median duration positive | Mean duration positive |
| --- | --- | --- | --- | --- | --- | --- | --- | --- |
| Wildtype | All | 75 | Model 1 | 0.085 ( 0.056 , 0.119 ) | 0.87 ( 0.72 , 0.96 ) |  | 8.1 ( 5.8 , 12.4 ) | 11.7 ( 8.4 , 17.9 ) |
| Alpha variant | All | 368 | Model 1 | 0.034 ( 0.025 , 0.043 ) | 0.91 ( 0.86 , 0.95 ) |  | 20.4 ( 16.1 , 27.4 ) | 29.5 ( 23.2 , 39.5 ) |
|  |  |  | Model 2 | 0.065 ( 0.047 , 0.085 ) | 0.94 ( 0.90 , 0.97 ) | 3.4 ( 2.2 , 4.2 ) | 14.0 ( 12.0 , 17.5 ) | 18.7 ( 15.6 , 24.1 ) |
| Alpha variant<br>(Ct matched to wildtype*) | Random subset 1 | 204 | Model 1 | 0.033 ( 0.020 , 0.047 ) | 0.88 ( 0.81 , 0.94 ) |  | 20.9 ( 14.8 , 34.1 ) | 30.1 ( 21.3 , 49.2 ) |
|  |  |  | Model 2 | 0.072 ( 0.045 , 0.102 ) | 0.93 ( 0.87 , 0.97 ) | 3.6 ( 2.0 , 4.4 ) | 13.2 ( 10.8 , 18.1 ) | 17.5 ( 13.9 , 24.9 ) |
|  | Random subset 2 | 204 | Model 1 | 0.037 ( 0.025 , 0.050 ) | 0.90 ( 0.84 , 0.95 ) |  | 18.9 ( 14.0 , 27.9 ) | 27.3 ( 20.1 , 40.2 ) |
|  |  |  | Model 2 | 0.065 ( 0.040 , 0.093 ) | 0.94 ( 0.88 , 0.97 ) | 3.2 ( 1.1 , 4.2 ) | 13.8 ( 11.2 , 19.0 ) | 18.5 ( 14.6 , 26.5 ) |
|  | Random subset 3 | 204 | Model 1 | 0.040 ( 0.029 , 0.053 ) | 0.93 ( 0.88 , 0.97 ) |  | 17.3 ( 13.2 , 24.2 ) | 25.0 ( 19.0 , 35.0 ) |
|  |  |  | Model 2 | 0.066 ( 0.043 , 0.094 ) | 0.95 ( 0.91 , 0.98 ) | 3.0 ( 1.0 , 4.1 ) | 13.5 ( 11.1 , 17.9 ) | 18.1 ( 14.4 , 24.9 ) |

Parameters shown are the exponential decay rate (k), initial proportion of positive individuals detected (Sensitivity proxy) and for Model 2 the additional time delay parameter.
Values estimated from parameters are median duration positive, calculated as $\log(2)/k$ with the time delay added on for Model 2, and mean duration positive, calculated as $1/k$ with the time delay added on for Model 2
\* Subset of Alpha samples randomly selected so that proportion of samples with initial N-gene Ct<24.5, 24.5<Ct<32.5 and Ct>32.5 matches the proportions seen in the wildtype samples

**Supplementary Figure 1:**
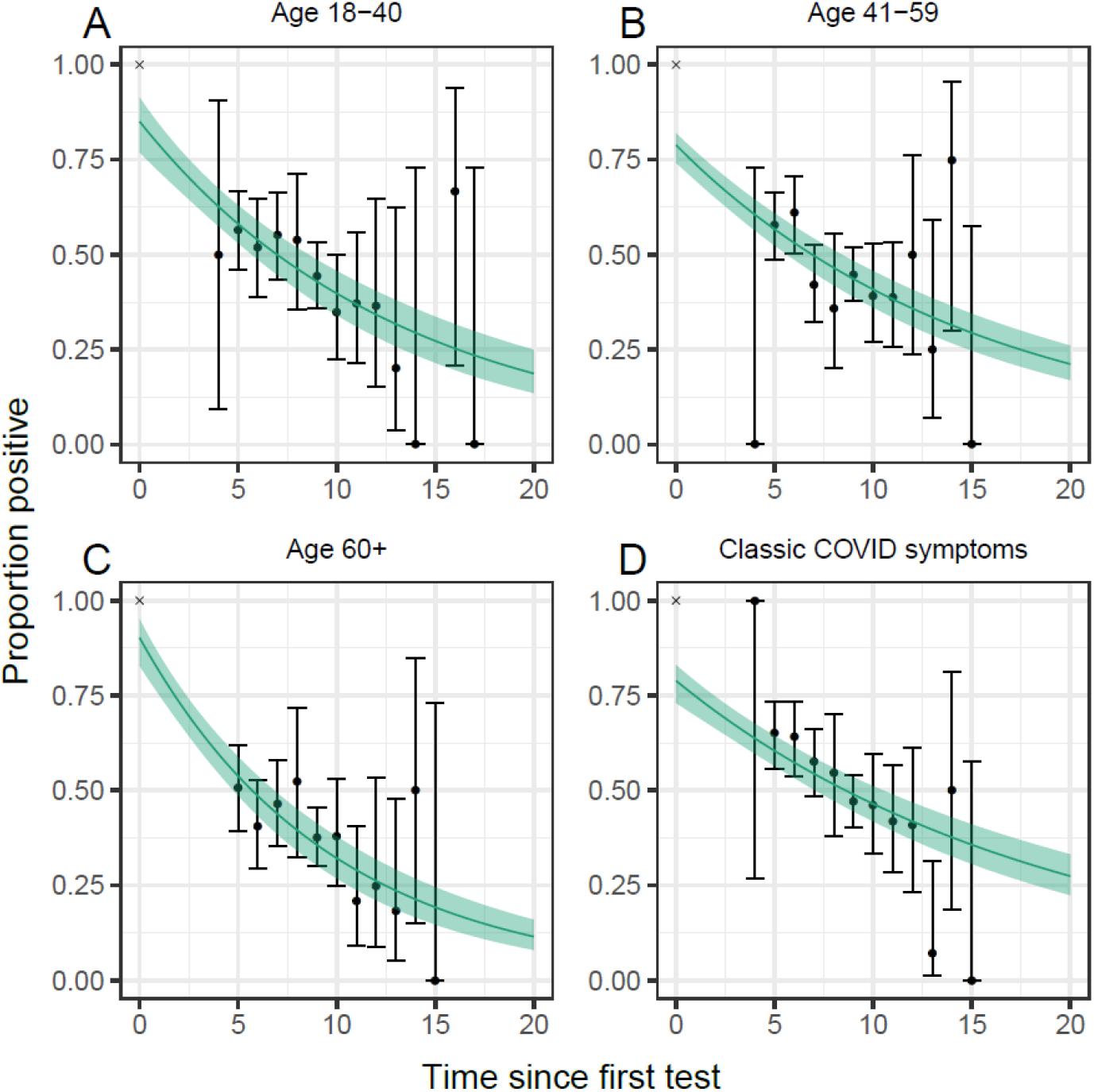
Best fit exponential decay model with no plateau (dark green line) and 95% confidence interval fit to subsets of the data. Data (black points) and 95% Binomial confidence intervals (black error bars) are included for the proportion of tests that were positive. (A) Model fit to all individuals aged between 18 and 40 years inclusive. (B) Model fit to all individuals aged between 41 and 59 years inclusive.(C) Model fit to all individuals aged 60 years and over. (D) Model fit to all individuals reporting having the most predictive COVID-19 symptoms in the month prior to their first test. Note that the models are not fit to the data as presented, but to the exact ordering of test outcomes for each individual.

